# Counterfactual Policy Learning for Personalized Treatment Using SGLT2 Inhibitors

**DOI:** 10.64898/2026.10.15.26365465

**Authors:** Dániel Sándor, Ákos Németh, Bálint Daróczy, Ferenc Sztanek, Mariann Harangi, Péter Antal

## Abstract

**Background:** Personalized medicine aims to tailor treatments to individual patients, but its implementation is limited by scarce clinical resources and heterogeneous treatment effects. Electronic health records now offer longitudinal data that can support data-driven therapy selection. We focus on SGLT2 inhibitors—medications with broad metabolic and cardiovascular benefits—whose clinical impact varies significantly based on individual patient profiles.

**Methods:** We propose a counterfactual Structural Causal Model (SCM) to estimate individual treatment effects (ITE) of SGLT2 inhibitors from observational data. The model assumes counterfactual stability and uses a Gumbel-Max sampling mechanism to capture stochastic treatment assignment while enabling differentiable inference. Optimal treatments are defined by maximizing the counterfactual value of future diagnostic outcomes.

**Results:** We evaluate the Counterfactual Gumbel-Max SCM using a doubly robust value estimator and show that, across diagnostic categories, it outperforms real-world prescriptions in predicted treatment value, highlighting subpopulations where SGLT2 inhibitors yield superior outcomes.

**Conclusion:** By integrating causal inference with electronic health data, this framework provides individualized counterfactual reasoning about SGLT2 treatment effects and can support clinicians in identifying optimal therapies for type 2 diabetes and comorbid conditions.

## 1 Background

Personalized medicine is gaining attention as more comprehensive individual-level health data become available throughout the treatment process. Modern Hospital Information Systems (HIS) provide rich longitudinal patient records, enabling physicians to make more informed clinical decisions based on detailed patient histories. However, the sheer volume and complexity of such data pose significant challenges for decision-making. In this context, machine learning models can play a crucial role by assisting clinicians in navigating complex information to identify optimal treatment strategies.

Machine learning models serve a dual purpose in personalized medicine. At a basic level, they can support clinical decision-making by recommending optimal treatments tailored to individual patient characteristics. More importantly, they enable modeling of the downstream effects of interventions, capturing how treatments influence patient trajectories over time.

By accurately modeling these downstream effects, it becomes possible to simulate treatment sequences and design optimal treatment policies involving multiple, potentially sequential interventions. Counterfactual modeling is a powerful approach that allows us to imagine hypothetical scenarios—asking “what if” questions about alternative treatment decisions—and thus learn from observational data where the intervention differs from what actually occurred.

A principled way to perform counterfactual reasoning is through Structural Causal Models (SCMs), which explicitly encode causal relationships between treatments, patient covariates, and outcomes. SCMs provide a formal framework to answer counterfactual queries, enabling personalized policy learning by simulating how different treatment choices would affect individual patients. Working from purely observational HIS data, we assume counterfactual stability [1] to uniquely identify treatment effects—thereby restricting the model to a single outcome by constraining the exogenous noise in the observations. We utilize an SCM to generate counterfactual data for policy training and explainable AI insights.

### 1.1 Counterfactuals in medical treatment

In medical practice, retrospective evaluation is vital for improving clinical decision-making. When a patient’s condition deteriorates following a specific intervention, we must ask: *Would the outcome have been different under an alternative treatment?* Answering this requires estimating a counterfactual scenario—a hypothetical ”parallel universe” where the treatment is toggled while all underlying exogenous factors remain constant. The hypothetical parallel treatment process is shown in Fig. 1.

**Fig. 1.**
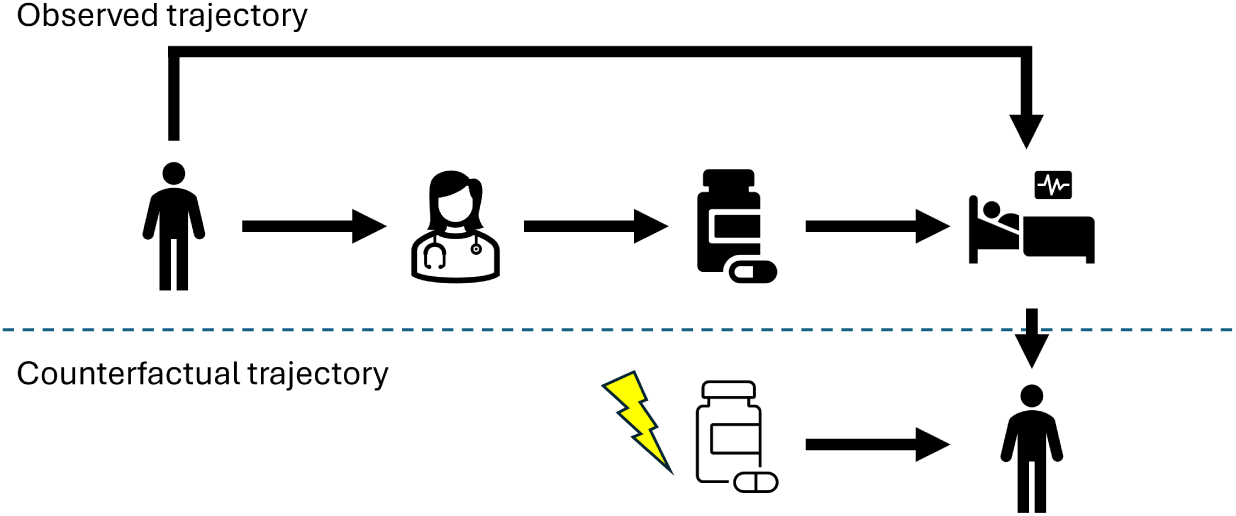
Counterfactual estimation: We observe a treatment process. Based on the outcome, we model the hypothetical scenario: What would have happened if the patient had been treated differently?

By comparing the factual outcome with the counterfactual state, we can determine the efficacy of our choice. If the counterfactual outcome is superior, it suggests a sub-optimal treatment path, providing a learning signal for future clinical scenarios. Although non-causal methods for treatment selection exist [2, 3], identifying the specific patient features that drive these differences enables better-informed treatment selection. To answer causal questions, we can create a causal model based on domain knowledge, observational, and interventional data [4].

In this work, we assume counterfactual stability to identify these latent scenarios. We utilize a Structural Causal Model (SCM) to generate counterfactual data, which are subsequently used for policy training and to generate explainable AI (XAI) insights regarding treatment selection.

Structural Causal Models (SCMs) provide a framework for answering counter-factual questions; however, formulating such a model requires specific identifiability assumptions to learn from observational data. In our case, the data sourced from Hospital Information Systems (HIS) is purely observational. While SCM parameters can be estimated from such data, exactly identifying a counterfactual model requires the assumption of counterfactual stability [1], which serves as a generalization of monotonicity within counterfactual SCMs.

Identifiability in SCM parameterization implies that we can uniquely determine the outcome of an intervention. Because multiple parameterizations can typically fit a given observational dataset, we assume counterfactual stability to restrict the model to a single outcome by constraining the exogenous noise of the observations.

Using a trained SCM, we aim to characterize the treatment policies currently used by clinicians and evaluate their performance. We specifically examine the treatment of type 2 diabetes (T2DM) with SGLT2 inhibitors. As these are relatively new medications, no universal standard of care has yet been established for their administration. Consequently, we seek to model current prescription patterns and derive an alternative treatment policy that is superior in terms of Aggregate Treatment Effect, as measured by a neutral Doubly Robust Outcome Estimator.

### 1.2 SGLT2 inhibitors

Sodium–glucose cotransporter-2 (SGLT2) inhibitors have become a central component of cardiometabolic and renal protection in T2DM, with large outcome trials demonstrating class-consistent reductions in heart-failure hospitalization and slowing of chronic kidney disease progression [5–10]. Treatment allocation in routine practice rarely follows a single decision rule, but rather reflects a multidimensional clinical assessment integrating metabolic control, renal function, cardiovascular phenotype, and comorbidity burden. The treatment policy analyzed in this study should therefore be interpreted as reflecting clinician behavior operating within established cardiometabolic safety boundaries, even though our modeling framework does not explicitly encode all contraindication rules at the algorithmic level.

In Hungary, these agents were introduced around 2015 and are now widely prescribed for patients with T2DM, particularly in the presence of cardiovascular or renal risk factors. Large cardiovascular and renal outcome trials have demonstrated consistent reductions in heart-failure hospitalization and slowing of chronic kidney disease progression across the drug class, with favourable effects on cardiovascular risk in selected populations [11–13].

Empagliflozin, dapagliflozin and ertugliflozin are the most commonly prescribed agents in Hungary. Although individual trials have highlighted specific clinical contexts, the therapeutic benefits of SGLT2 inhibitors are generally interpreted as largely class-consistent effects, with differences between the pharmaceuticals often reflecting variations in study populations and endpoint definitions rather than fundamentally distinct pharmacological mechanisms [5, 8, 14].

However, treatment allocation in routine clinical practice rarely follows a single decision rule. Conversely, the prescription of behavior reflects a multidimensional clinical assessment integrating metabolic control, renal function, cardiovascular phenotype, and comorbidity burden. Observational electronic health record data, therefore, provide an opportunity to investigate how such complex treatment decisions emerge in real-world practice and whether alternative treatment policies may be associated with improved predicted clinical trajectories.

Over the past decade, sodium–glucose cotransporter-2 (SGLT2) inhibitors have evolved from glucose-lowering agents to a central component of cardiometabolic and renal protection in T2DM. Large cardiovascular outcome trials [5–7], have demonstrated consistent reductions in heart-failure hospitalization and favourable cardiovascular risk profiles across multiple agents. Subsequent studies [8–10] have further substantiated the significance of SGLT2 inhibition in the management of heart failure and the progression of chronic kidney disease. Consequently, therapeutic decision-making regarding SGLT2 inhibitors is increasingly influenced by an integrated cardiometabolic and renal risk framework rather than glycaemic control alone. Accordingly, the treatment policy analyzed in this study should be interpreted as reflecting clinician behavior operating within these safety boundaries, even though the present modeling framework does not explicitly encode all contraindication rules at the algorithmic level.

### 1.3 Related work

#### 1.3.1 Counterfactual inference

Counterfactuals are the third rung of causality after observations and interventions [15]. They address the question of what would have happened under a different intervention if we have already observed the outcome of a different one. In the medical domain, randomization is not always feasible, but we can still draw conclusions about causal effect estimation [16] based on observational data. Authors in [17] propose a joint treatment-outcome model and handle dependencies between events in patient records. To train a treatment policy on observational data only, we need to know the outcomes of interventions other than the observed ones. To tackle this, Buesing et al. [18] proposed Counterfactually-Guided Policy Search (CF-GPS). They showed that SCM-based counterfactual data can debias the learning process and improve on existing models. One of the leading problems is the identification of the counterfactuals. In the continuous case it is solved by De Brouwer [19] and in the discrete categorical case by Oberst and Sontag [1]. To estimate individual-level treatment effects (ITE), Shalit et al. [20] derived a family of algorithms. Another approach for estimating ITE, proposed by Yao et al. [21], is based on local similarity preservation. When confounders are present, we can apply the work of Louizos et al. [22] to obtain meaningful ITE estimates.

#### 1.3.2 Treatment with SGLT2 inhibitors

SGLT2 inhibitors have been used to manage T2DM since the mid-2010s, and are now widely recognized as cardiometabolic therapies, with benefits extending beyond glycaemic control. A substantial body of literature provides clinical recommendations that summarize best practices for their use [23]. However, real-world uptake remains heterogeneous across healthcare systems. Despite the existence of substantial evidence that demonstrates the cardiovascular and renal protective effects of SGLT2 inhibitors, multiple studies suggest that this class of medication has not been utilized to its full potential in eligible patient populations [24–26].

In routine clinical practice, treatment allocation rarely follows a single decision rule. Instead, physicians must consider a multitude of clinical factors, including metabolic control, renal function, cardiovascular risk profile, and comorbidity burden, when determining whether to initiate SGLT2 inhibitor therapy. This inherent complexity makes treatment selection a suitable candidate for data-driven modeling approaches aimed at characterizing or optimizing treatment policies. A number of statistical and machine learning approaches have been proposed to support treatment selection in T2DM. Cardoso et al. developed a phenotype-based algorithm focusing on differential HbA1c responses using regression modeling [27]. The treatment selection framework developed by Dennis et al. is based on the principle of Bayesian causal forests [28]. A similar approach was adopted in other studies, which explored counterfactual prediction models for SGLT2 inhibitor effects on renal outcomes [29] and time-dependent modeling of treatment trajectories in observational datasets [30, 31]

### 1.4 Overview

We use data from real patients with diabetes and train an SCM to examine how treatment affects diagnostic variables and the diagnosis itself. We use diagnostic probabilities as outcomes to quantify improvement across diabetes and 10 selected comorbidities. We calculate the best potential outcome for every treatment to propose a treatment policy. To test our models, we use the observed medication policy of the test dataset and models trained exclusively on observational data. We show that the models are not only superior in terms of diagnostic probabilities but also in Disability-Adjusted Life Years (DALYs). This way, we also quantify the effects of treatments at the population level. Testing the SCM policy against real-world interventions is itself a counterfactual problem; to address it, we use Doubly Robust Outcome Estimation. The overview of the training and evaluation pipeline is seen in Fig. 2.

**Fig. 2.**
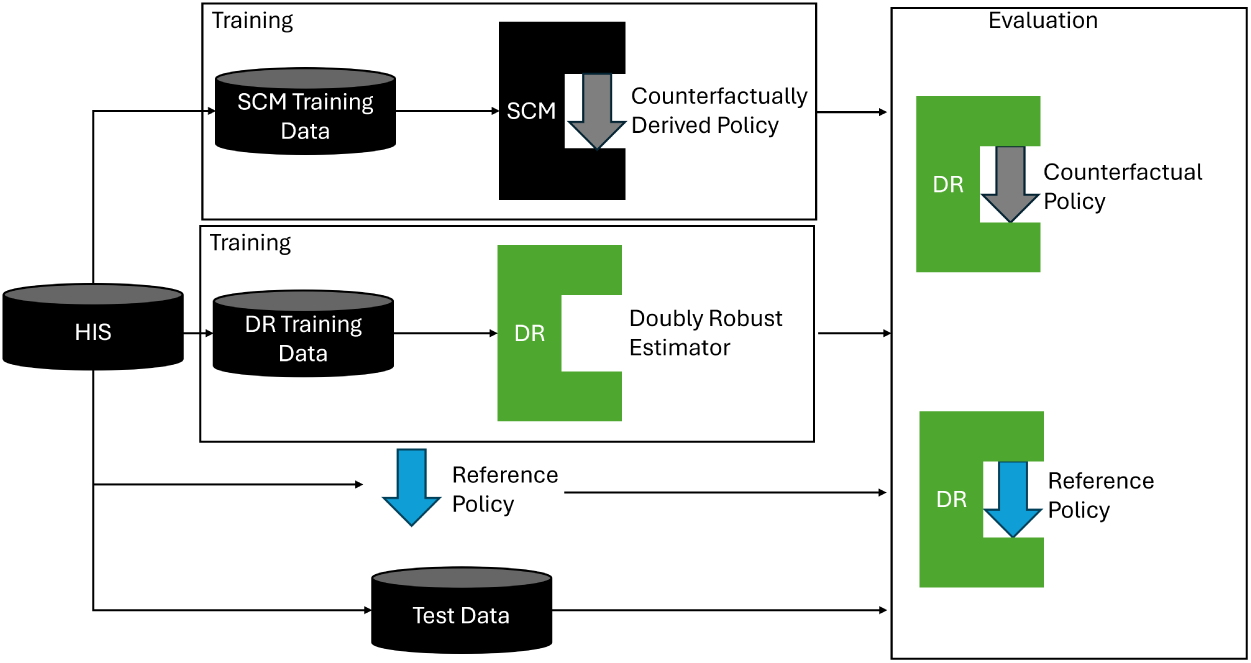
Overview of the modeling process.

Our contributions are the following:

- We train an SCM on real-world patient data to model treatment.
- We use the SCM to characterize the treatment policy in use by doctors.
- We generate counterfactual data to train medical treatment policies.
- We propose an evaluation strategy for policies that measures ITE across multiple treatment types.

The paper is structured as follows: Section 2 describes the models and data structures used in Causal inference and the calculation of the Counterfactual policy. In Section 3 we describe the quantitative results achieved by the Counterfactual SCM-based Policy and investigate the treatment policy in use by doctors. In Section 4 we discuss the strengths and limitations of our models and the possible alternatives. Finally, in Section 5 we end with a conclusion and possible future directions for our models.

## 2 Methods

We model the Partially Observed Markov Decision Process (POMDP) with a Gumbel-Max SCM parametrized by a set of neural networks. We first construct and evaluate the SCM, then use it to train a policy network optimized for the chosen treatment duration. We present the results of the policy network trained for single-treatment prediction. This means that the policy net predicts the medication to be used and the diagnosis is evaluated based on the patient’s next case in the treatment history.

We use the Gumbel-Max SCM described by Oberst and Sontag [1], but our goal is not policy evaluation but to enable counterfactual policy learning. For this reason, we first construct the SCM and train it to learn the parameters. After this we learn a treatment policy with a simple neural network and use the counterfactual predictions to train the policy network with reinforcement learning. Our goal with the training is to maximize the Individual Treatment Effect (ITE).

### 2.1 Gumbel-Max SCM

In accordance with the notation of Oberst and Sontag [1], an SCM *M* is a set of unobserved **U** = *{U*_1_*, …, U_n_}* and observed **X** = *{X*_1_*, …, X_n_}* variables and functions **f** = *{f*_1_*, .., f_n_}*. We use the SCM to model one treatment step in the treatment policy. Thus, we add the temporal element to the variables denoted by **X**(**t**).

We classify the observed variables into observations **LAB**(**t**) = *{LAB*_1_(*t*)*, …, LAB_n_*(*t*)*}*, diagnosis **DIAG**(**t**) = *{DIAG*_1_(*t*)*, …, DIAG_n_*(*t*)*}* and treatment *T* (*t*). The corresponding unobserved variables are denoted by **U_L_**(**t**), **U_D_**(**t**)*, U_T_* (*t*).

Our core assumptions about dependencies in the causal Directed Acyclic Graph (DAG) *G* are as follows. Every lab influences the diagnosis of every disease in a given timestep *t*: *LAB_i_*(*t*) *→ DIAG_j_*(*t*)*∀i, j*, every observation is dependent on the previous observations and the treatment only (they are independent of all previous diagnoses given the previous observation and treatment) *LAB_i_*(*t*) *→ LAB_j_*(*t*)*∀i, j*; *T* (*t*) *→ LAB_j_*(*t*)*∀j*. Treatment in every time step *t* is dependent on both the observation and the diagnoses *LAB_i_*(*t*) *→ T* (*t*)*∀i*; *DIAG_i_*(*t*) *→ T* (*t*)*∀i*. The resulting *G* is shown in Fig. 3.

**Fig. 3.**
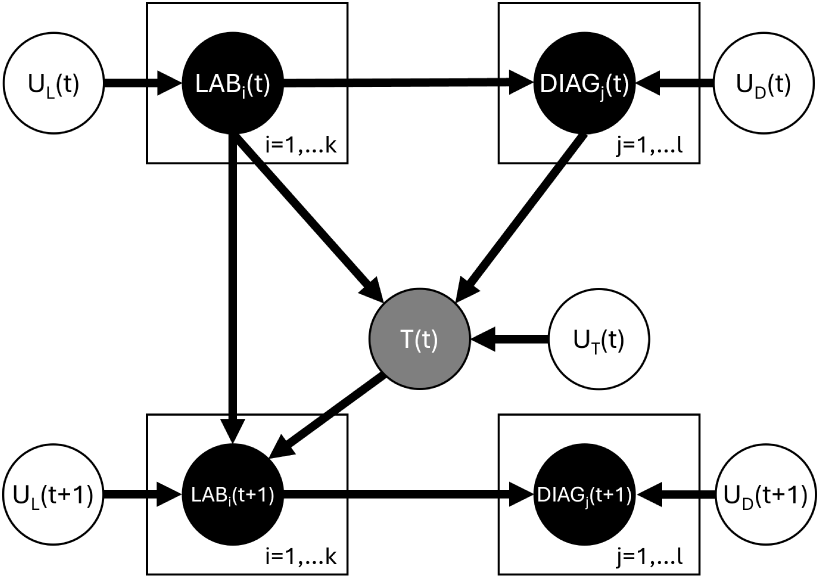
SCM of treatment in time step *t*.

To identify counterfactual treatment effects, we assume counterfactual stability (granted by the Gumbel-Max SCM). We want to maximize the reward with a given treatment policy *π*^. This means that after the treatment *T ^′^*(*t*) is assigned, we use the SCM to predict *P* (*LAB_i_*(*t* + 1)*|T* (*t*)*, T ^′^*(*t*), **LAB**(**t**)). As we assume additive Gumbel noise, we can think of the observed output as

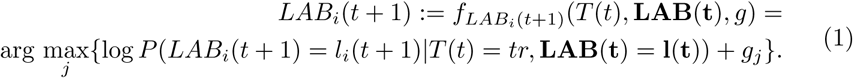

This ensures counterfactual stability. To sample *g*, we use the Monte Carlo method described by Oberst and Sontag [1]. To summarize the counterfactual prediction, we first sample *g* from

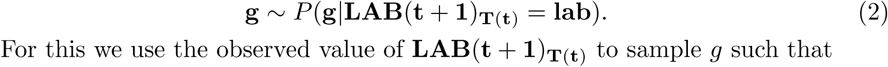

For this we use the observed value of **LAB**(**t** + **1**)**_T_**_(**t**)_ to sample *g* such that

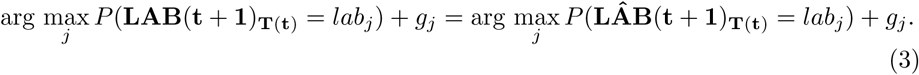

From this, we formulate the counterfactual prediction like

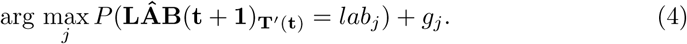

As the diagnoses are conditionally independent of the treatment given the observations, we can simply predict the *DIAG_i_*(*t* + 1) from the counterfactual **LAB**(**t** + **1**).

Once the SCM is constructed, we need to identify a policy. We do this by predicting the counterfactual outcomes for a previously unseen ”policy training set”. This way, we can predict the usefulness of each treatment with the SCM having access to the observed outcomes. The policy is trained on counterfactual rewards generated by the SCM; it does not have direct access to the outcomes. After we predicted outcomes with the SCM, we trained the policy model to predict outcomes for each value of the treatment variable. Further information on deriving the policy from the counterfactuals can be found in A and on the hyperparameters in B.

### 2.2 Evaluation

We evaluate the results of our policy by off-policy evaluation using Doubly Robust (DR) estimation. To do this, we train a random forest model as an outcome predictor *µ*, and we use multinomial regression for propensity score estimation *p*^(*T_iπOBS_ | X_i_*). Both the outcome estimator and the propensity score estimator are trained on the DR training set, and all the evaluation is done on the DR test set.

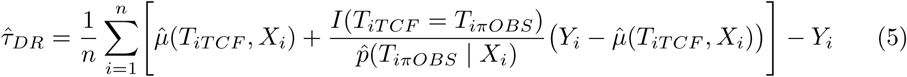

### 2.3 Data

We use real data collected in Hungarian hospitals from 2000 to 2021. Data collection was conducted in the Northern Great Plains region at the University of Debrecen Clinical Center and the Szabolcs-Szatmár-Bereg County Hospitals. The entire dataset contains more than 31000 cases from 6100 patients. We use blood tests, BMI, blood pressure and demographic data to train the policy model. The data is discretized to enable discrete learning of the SCM outcomes. The dataset was partitioned into three disjoint parts based on patient IDs, ensuring that each patient belongs to exactly one model’s training set. The three subsets were also partitioned into training and test sets. The ratios used for the different models are: 0.4 for DR estimation and 0.3 for the SCM and the policy network. In each subgroup, 30% of patients were used only for evaluation.

## 3 Results

We demonstrate that the proposed counterfactual policy models effectively match the performance of the observed behavior policy (physician treatments) for diagnostic outcomes and, in several instances, significantly outperform it. To isolate the impact of the SCM-generated counterfactual data, we conduct an ablation study comparing our approach against models with identical architectures trained solely on observational data. The results indicate that policies trained with counterfactual augmentation achieve superior outcomes across most tested comorbidities. Furthermore, we utilize the SCM to characterize existing clinical policies by estimating their expected long-term outcomes and identifying divergence points where alternative counterfactual treatments suggest improved patient trajectories.

### 3.1 Treatment effects

To see how much improvement is expected we first show the average of true outcomes of diagnosis probabilities across diseases in Fig. 4. In this we already see that based on the choice of the cohort 50% of people are diagnosed with diabetes and other comorbidities are mostly present in 10-20% of the population.

**Fig. 4.**
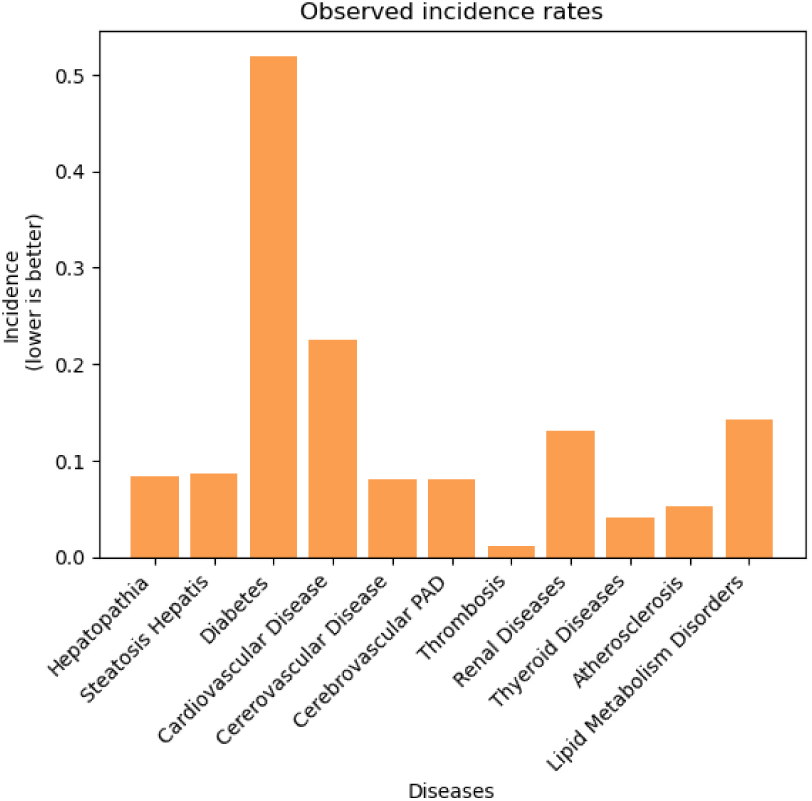
Observed disease diagnosis in the evaluated population.

#### 3.1.1 Improvement of the SCM

The counterfactual policy derived from the SCM matches or exceeds the performance of the observed clinical behavior policy across all investigated comorbidities. As illustrated in Fig. 5, we observe a minimum improvement of 1% in diagnostic outcomes for every comorbidity, with gains of 5–10% being frequent. Notably, diabetes progression is significantly mitigated under the SCM-derived policy. This serves as a clinical sanity check, confirming that the model captures the known efficacy of interventions such as SGLT2 inhibitors. The major improvements over the observed policy are expected, as the SCM has access to the observed treatment outcomes. However, it is important to note that this is a counterfactual treatment selection, which is only practical for post-hoc explanations. We show the performance of the policy models in section 3.1.2.

**Fig. 5.**
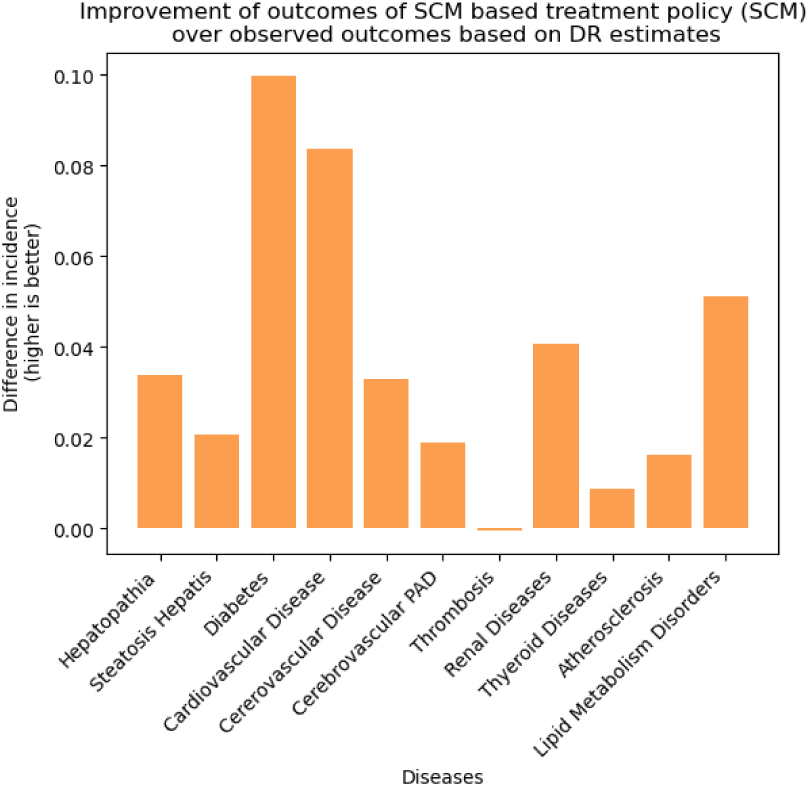
Improvement of counterfactual treatment policy over the observed policy of physicians.

#### 3.1.2 Improvement of suggested policies

The SCM can generate counterfactual training data, but it is not the ideal tool for predictive modeling. If we want to create a decision support system for medical professionals to use, the SCM is good for generating explanations and answering causal questions, but to recommend treatments, a traditional predictive model is better suited. Here, we analyze the performance of two such predictive models trained on the counterfactually generated datasets.

We use a simple feed-forward neural network and gradient boosting as models for outcome prediction. Our policy function always selects the action with the best average diagnosis outcomes. To see how much the counterfactually assisted models improve in predictive performance, we train models with comparable hyperparameters using solely observational data. Finally, we add a random policy to verify if the improvement made by the observational-only policies is significant.

For evaluation, we use bootstrapping on the doubly robust estimators and predict the results on the same test set in each iteration. We get confidence intervals for our predictions with all of the tested models. The mean outcomes predicted for all policies with DR estimation are shown in Fig. 6 and the mean improvement of the outcomes for the major disease groups is shown in Table 1.

**Fig. 6.**
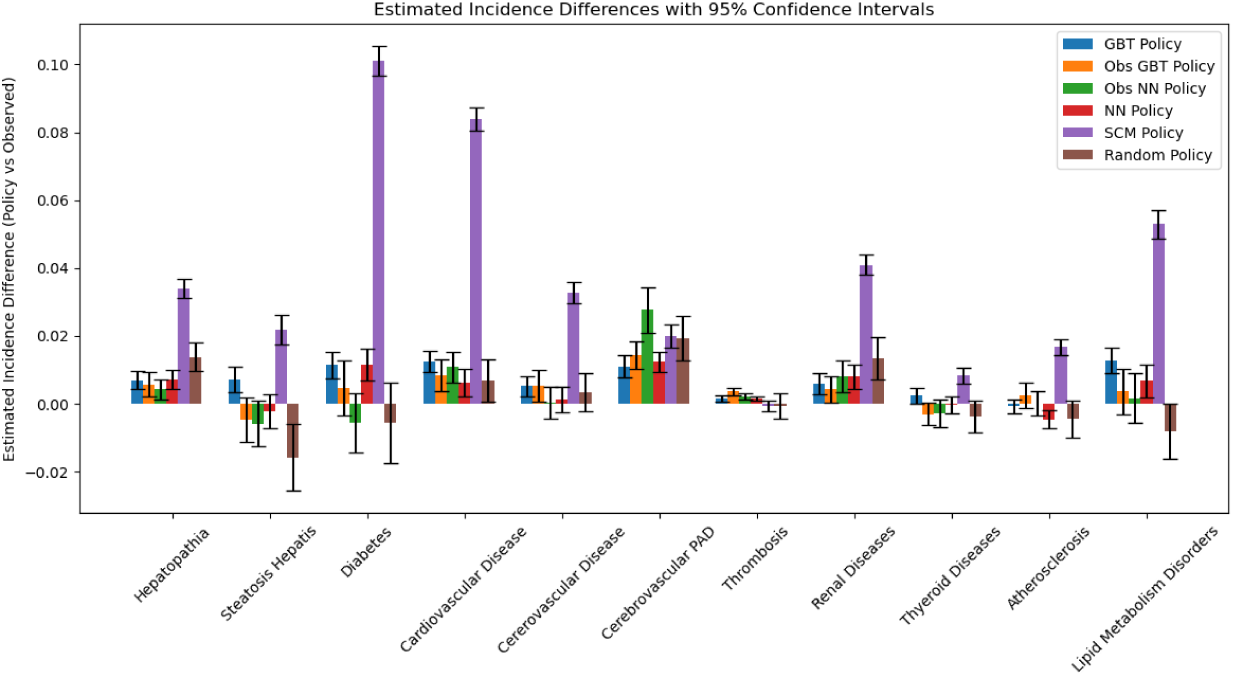
Comparison of policy improvements over observed policy. The differences are calculated by DR estimation (higher is better).

**Table 1.** Updated Mean DR Estimates for Major Disease Categories.

| Model | Hepato-pathia | Diabetes | Cardio-vascular Disease | Cerebro-vascular Disease | Renal Diseases | Thyroid Diseases | Lipid Metab. Disord. |
| --- | --- | --- | --- | --- | --- | --- | --- |
| GBT | 0.0080 | **0.0104** | **0.0127** | 0.0043 | 0.0048 | **0.0014** | 0.0116 |
| Obs GBT | 0.0056 | 0.0046 | 0.0083 | 0.0052 | 0.0042 | -0.0031 | 0.0036 |
| Obs NN | 0.0057 | -0.0098 | 0.0099 | 0.0054 | 0.0087 | -0.0047 | -0.0080 |
| NN | 0.0062 | 0.0089 | 0.0064 | **0.0057** | 0.0027 | -0.0000 | **0.0117** |
| Random | **0.0137** | -0.0057 | 0.0068 | 0.0035 | **0.0135** | -0.0037 | -0.0081 |
| **SCM** | **0.0330** | **0.1133** | **0.0869** | **0.0310** | **0.0458** | **0.0100** | **0.0515** |

We found that the SCM’s performance is superior to all other models, and access to the observed outcomes is a considerable advantage even for predicting counterfactual values. The models trained on counterfactual data outperform their counterparts trained solely on observational data across major disease groups. The observational models and the random policy both yield high-variance outcomes. For some diseases, observational models perform acceptably, but their deterioration on others makes them unfit for decision support. The NN and GBT models’ performance is similar; they perform well across different disease groups, suggesting an inherent difference in how they predict outcomes.

While most policy models yield improvements over the observed policy that remain too small to be considered clinically relevant, the SCM achieves gains that cross this threshold—demonstrating that clinically meaningful improvement is attainable when observed outcomes are available at training time. An important open question is how closely this clinically relevant performance can be approached by models that rely solely on observational data at inference time.

### 3.2 Understanding existing treatment policies

In clinical practice, SGLT2 inhibitors are primarily used in patients with major cardiovascular disease and chronic kidney disease. For patients with type 2 diabetes, the frontline treatment remains metformin-based therapies. If they have comorbidities such as kidney or heart disease, they are then considered for SGLT2 inhibitors. Regarding the differences between specific drugs, the two most prominently used are empagliflozin and dapagliflozin. These are considered comparable in effect, though empagliflozin is often prescribed to patients with heart disease due to a larger body of clinical studies in that population.

As demonstrated, the DR outcome estimation shows that the SCM can be a valuable tool for counterfactual estimation. In this subsection, we show that the SCM can pinpoint suboptimal transitions in the treatment process and serve as a valuable tool for estimating the effect of deviations from a superior policy. For this, we use the SCM’s counterfactual predictions on the test set to detect past mistakes and show potentially better outcomes. It is important to note that this functionality is not a predictive policy; we merely use it to explain existing policies.

As shown in Fig. 7 we can see that in most of the cases the SCM is in agreement with medical professionals however, it would prescribe more SGLT2 inhibitors overall. To see what factors the model considers, we can calculate the mutual information between its predictions and the input variables, as shown in Fig. 8. Based on this, the observed data suggests a focus on factors more closely related to T2DM, while the SCM decides based on factors indicative of the comorbidities as well, for example, Serum Cholesterol (CHOL) related to heart conditions and Creatinine (KREA) related to kidney function.

**Fig. 7.**
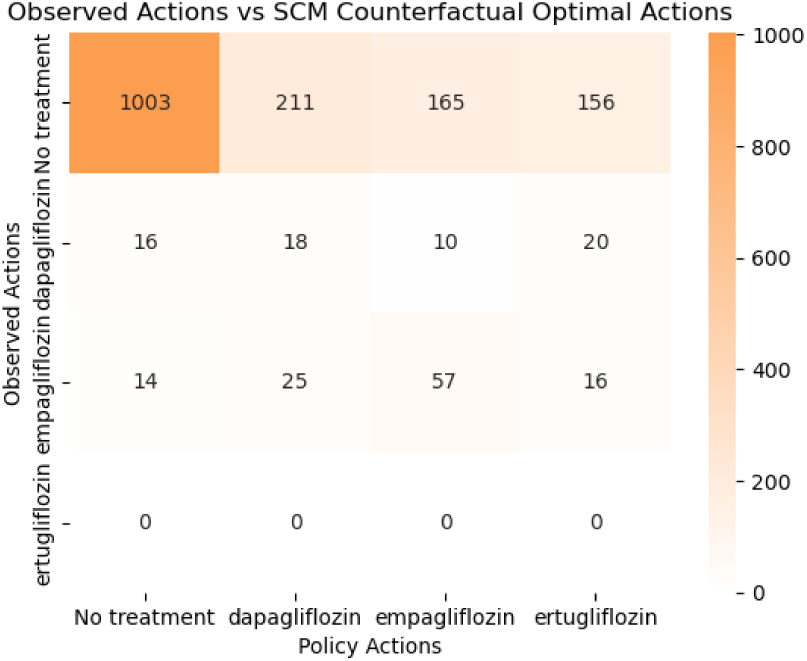
The treatments assigned to individuals by the SCM and by physicians.

**Fig. 8.**
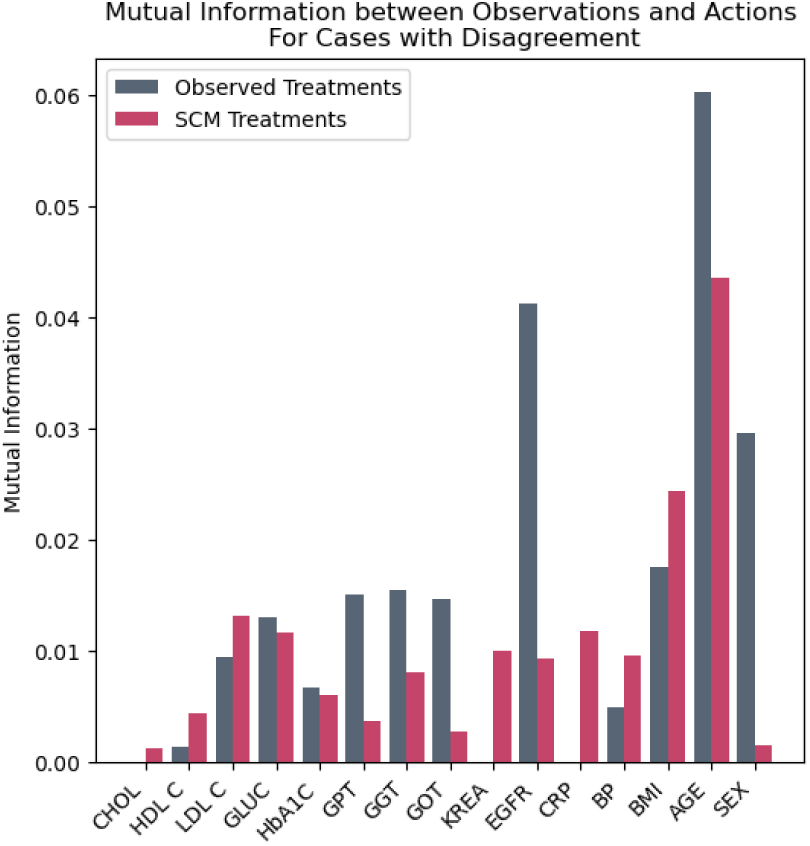
Mutual Information score of observed variables and predictions. The proposed method places the focuse more towards predictors of comorbidities (CHOL, KREA).

Further analysis of the disagreements shows that although the SCM tends to recommend more SGLT2 inhibitors overall, a large portion of these recommendations occurred before 2014, when medical practice had not yet adapted to them or, in most cases, they were not yet available. Table 2 shows the number of cases where the two policies align or conflict. We see that around 50% of conflicts date from before 2014, and another large portion fall into the ”Delayed Alignment” category, in which the patient was eventually prescribed an SGLT2 inhibitor.

**Table 2.**
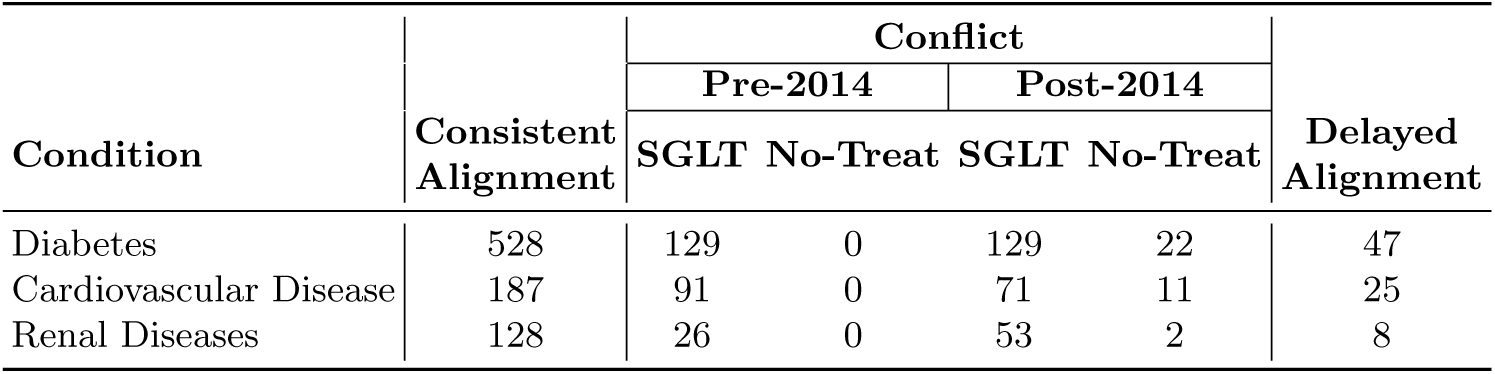
Comparative Alignment of SCM Recommendations vs. Observed Clinical Policy. The SCM identifies earlier eligibility for SGLT2 therapy; conflicts are primarily driven by SCM-led prescriptions prior to the 2014 policy shift. ”Delayed Alignment” highlights cases where observed policy eventually synchronized with SCM-predicted interventions.

### 3.3 Validation against the 2014 SGLT2 availability shift

As SGLT2 therapies became more widely available in 2014, a subset of patients was transitioned to these treatments. Our test set includes 24 such cases; in every instance, the SCM predicted the new SGLT2 therapy either before or at the same time it was prescribed. The treatment trajectories for these patients, including the initial SCM recommendations, are illustrated in Fig. 9.

**Fig. 9.**
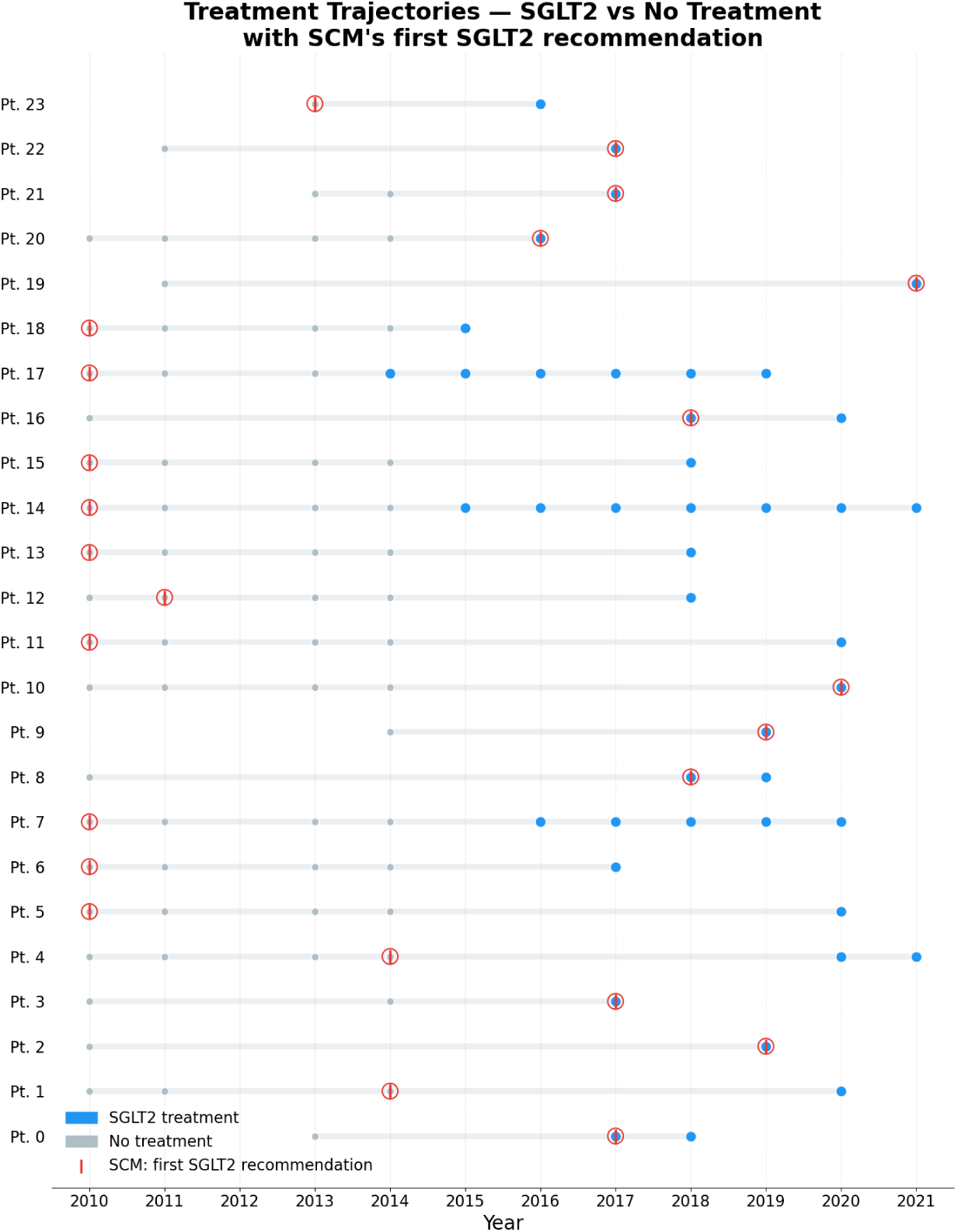
Treatment trajectories of patients in the test set for whom the treatment changed as SGLT2 inhibitors became available. We show that our model predicted they should receive SGLT2 inhibitors earlier or at the same time as their treatment started for every patient.

## 4 Discussion

Although we observe average improvements across all outcomes, these gains do not guarantee that disease progression improves for every individual patient. Deciding how to weigh competing outcomes against one another is a value-laden judgment that clinicians are often better placed to make than a machine learning model, so the system is best used under the supervision of medical professionals. Our policy currently selects the treatment that maximizes the average diagnosis probability across diseases, which implicitly weights all comorbidities equally; more principled objectives—for example, weighting outcomes by disease severity or DALY burden—may better reflect clinical priorities.

Further limitations include the positivity assumption, that all treatment options carry non-zero probability for each patient, which is partially violated in our dataset, since SGLT2 inhibitors were only available for selected tests before approximately 2014. We address this by reporting the temporal breakdown of policy conflicts in Table 2, but future work should consider restricting the primary causal analysis to post-2014 data where positivity is better supported.

Translating the current framework into a clinical decision support tool would require additional steps beyond what is demonstrated here. The policy network must be validated prospectively rather than through retrospective off-policy estimation; a randomized pilot study in which the tool’s recommendations are surfaced to clinicians alongside their usual information, without overriding clinical judgment, would be the appropriate next step.

### 4.1 Global Burden of Disease

Although the benefits of the improved policies are small in terms of diagnostic outcomes, they could provide a societal-level improvement. DALY [32] and other Global Burden of Disease (GBD) metrics measure how many years an individual loses in their lifetime because of a given disease, with disability taken into consideration as well. We show that improving the policy can substantially reduce DALY scores in a society. For this, we use the publicly available GBD [32] metrics to calculate the individual-level improvement. We aggregate at the level of the evaluation population. The results are seen in Table 3. Further information on our methodology for calculating the DALY scores can be found in C.

**Table 3.** Bootstrap DALY rate improvement (Observed - Policy) for 100000 people.

| Model | Mean Improvement |
| --- | --- |
| GBT | 76.9 |
| NN | 98.5 |
| SCM | 182.9 |

We see that a considerable number of life years can be saved by choosing the correct treatment with either policy. The NN model outperforms the GBT model, indicating it is a better policy with respect to DALY weighting. The SCM’s performance is still double that of the NN’s. This improvement indicates that beyond diagnostic outcomes the impact of the diseases can still be considered and medically sound performances can still be reached.

## 5 Conclusion

We have created a counterfactual modeling tool for estimating treatment effects. Based on this SCM, we have generated a counterfactual training dataset. We trained our own treatment policies that matched the performance of the observed policies used by medical professionals and outperformed them in some cases. Based on the SCM, we can understand better how we treat patients, and we can improve our existing policies for a more effective treatment process. We found that some key lab values are often overlooked when choosing treatment. Based on the incorporation of these the treatment selection process might be improved further.

Although the method still has limitations, it presents a desirable causal direction in medical diagnostics. Since predictions are causally explainable, we can use this tool to simulate disease progression trajectories over longer time periods as well, with minimal extensions. This leads to better cooperation with medical professionals and hopefully gives them a tool to use in a highly important but less-understood field.

## Data Availability

The datasets generated and/or analysed during the current study are not publicly available due to the terms of the ethical approvals governing patient-derived hospital data (TUKEB 44549-2/2018/EKU; DE RKEB/IKEB 6697-2024), but are available from the corresponding author on reasonable request.

## List of abbreviations

SGLT2: Sodium–Glucose Cotransporter-2 (Inhibitors)
SCM: Structural Causal Model
T2DM: Type 2 diabetes
ITE: Individual Treatment Effects
HIS: Hospital Information Systems
CF-GPS: Counterfactually-Guided Policy Search
HbA1c: Hemoglobin A1c (Glycated Hemoglobin)
DALY: Disability Adjusted Life Years
GBD: Global Burden of Disease
ATE: Aggregate Treatment Effect
POMDP: Partially Observed Markov Decision Process
DAG: Directed Acyclic Graph
DR: Doubly Robust (Estimation)
ReLU: Rectified Linear Unit (Activation Function)
GBT: eXtreme Gradient Boosting
NN: Neural Network

## Declarations

### Ethics approval and consent to participate

This study did not involve any experimental intervention on human participants; it is a retrospective analysis of anonymized clinical records collected as a by-product of routine care between 2000 and 2021. The study was approved by the Scientific and Research Ethics Committee of the Hungarian Medical Research Council (TUKEB; approval number 44549-2/2018/EKU) and by the Regional and Institutional Research Ethics Committee of the University of Debrecen (DE RKEB/IKEB; approval number 6697-2024), as part of the University of Debrecen Clinical Center’s data mining and machine learning initiative. All methods were carried out in accordance with the relevant guidelines and regulations and with local legislation and institutional requirements. Informed consent was waived by the above-named ethics committees, in accordance with national legislation and institutional requirements, because the study used anonymized data derived from routine clinical care.

## Clinical trial number

Not applicable.

## Consent for publication

Not applicable.

## Availability of data and materials

The source code generated for this study is available on: https://github.com/sandordani/Counterfactual-SGLT2-Policy.

## Authors’ contributions

DS: Conceptualization, Methodology, Software, Validation, Formal analysis, Investigation, Writing - Original Draft. ÁN: Conceptualization, Validation, Resources, Data Curation, Writing - Review & Editing. BD: Conceptualization, Methodology, Validation, Resources, Writing - Review & Editing. FS: Validation, Resources, Writing - Review & Editing. MH: Validation, Writing - Review & Editing. PA: Conceptualization, Methodology, Formal analysis, Investigation, Resources, Writing - Review & Editing.

## Acknowledgements

Not applicable.

## Appendix A Deriving the policy

To formulate a treatment policy, we use observational values of **LAB**(**t**) and **DIAG**(**t**) to predict treatment variables using a policy model. To train the policy model, we first generate counterfactual samples for each possible treatment for every sample in the policy training set, using the SCM. Using the counterfactual samples, we train the policy model for the outcome-estimation task. Thus, the policy model learns to predict **DIAG**(**t** + **1**) from only the observed inputs. Finally, we can use simple strategies to calculate the best outcome, according to the policy model. An overview image of the policy generation in the context of the SCM is included in Additional file 1: Figure S1.

In most cases we cannot say that trajectories are superior to each other, instead one policy may perform better on disease A while another performs better on disease B. For now, we use the average of the outcome probabilities for each action as the measure of the policies’ performance to obtain the treatment. However, in the future, it is worth considering aggregating the disease outcomes in a more principled way.

To test the model’s sensitivity to structure, we propose an eXtreme Gradient Boosting (GBT) and a Neural Network-based approach to learn treatment outcomes. We train these models on the policy train set, augmented with the SCM’s counterfactual predictions, to ensure sufficient data to model each treatment’s outcomes.

## Appendix B Model parameters

We discuss the hyperparameters of the models used for the SCM parameters, policies and evaluations.

### B.1 SCM parameters

The SCM is parametrized by neural networks learning

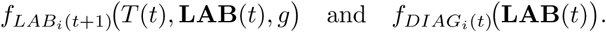

The networks are feedforward, consisting of two layers of 128 and 64 neurons, with ReLU activations in the hidden layers and soft-max on the output. For the networks that parameterize the diagnosis functions *f_DIAGi_*_(*t*)_(**LAB**(**t**)), the outputs are sigmoids representing the Bernoulli parameters for each disease. The neural networks are classifiers predicting low, normal, or high categories for tests and a probability between 0 and 1 for disease diagnosis outcomes. We use a cross-entropy loss for training, and each model is trained for 30 epochs.

### B.2 Doubly Robust Estimator

The doubly robust estimator consists of an outcome model and a propensity model. The outcome model is a Random Forest Classifier for each disease diagnosis; each classifier contains 200 estimators. The propensity model is a Multinomial Logistic Regression model with 1000 maximum iterations and L2 regularization.

### B.3 Extreme Gradient Boosting

The GBT model is parameterized with 500 estimators, a maximum depth of 6, and a learning rate of 0.1 for each outcome. They are jointly trained using sklearn’s Multi-outputRegressor, whereas in the observational case, 4 separate models are trained for the outcomes of 4 different treatments.

### B.4 Policy Neural Network

The policy NN is a feed-forward network with hidden layers of sizes 128, 64, 32. ReLU activations and the outcomes are predicted with a linear activation. The network is trained for 20 epochs with Adam optimizer.

## Appendix C DALY methodology

For each disease category tracked in our model, we extracted the age- and sex-standardized DALY rate per 100,000 population for Hungary. The policy-driven reduction in disease incidence probability — estimated via the doubly robust estimator for each model — was multiplied by the corresponding DALY rate and summed across all disease categories. Formally, for a cohort of *N* patients, the estimated DALY improvement is:

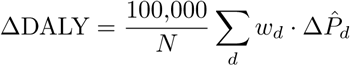

where *w_d_* is the GBD DALY rate for disease *d*, and Δ*P*^^^*_d_* is the doubly robust estimate of the reduction in incidence probability for disease *d* under the evaluated policy relative to the observed policy. Confidence intervals were obtained by applying the same formula to each bootstrap replicate of the DR estimator.

